# The new Coronavirus (SARS-CoV-2) in Central America: Demographic-spatial simulations, Analyses of Molecular Variance (AMOVA) and Neutrality Tests in complete genomes from Belize, Guatemala, Cuba, Jamaica and Puerto Rico

**DOI:** 10.1101/2020.12.26.20248872

**Authors:** Pierre Teodosio Felix, Robson da Silva Ramos, Dallynne Bárbara Ramos Venâncio, Eduarda Doralice Alves Braz Da Silva, Rosane Maria de Albuquerque

## Abstract

In this work, we evaluated the levels of genetic diversity in 38 complete genomes of SARS-CoV-2 from five Central American countries (Belize, Guatemala, Cuba, Jamaica and Puerto Rico) with 04, 10, 2, 8 and 14 haplotypes, respectively, with an extension of up to 29,885 bp. All sequences were publicly available on the National Biotechnology Information Center (NCBI) platform. Using specific methodologies for paired F_ST_, AMOVA, mismatch, demographic-spatial expansion, molecular diversity and for the time of evolutionary divergence, it was possible to notice that only 79 sites remained conserved and that the high number of polymorphisms found helped to establish a clear pattern of genetic non-structuring, based on the time of divergence between the groups. The analyses also showed that significant evolutionary divergences within and between the five countries corroborate the fact that possible rapid and silent mutations are responsible for the increase in genetic variability of the Virus, a fact that would hinder the work with molecular targets for vaccines and medications in general.

## 1. Introduction

COVID-19 is the acute respiratory infection that has been causing a great impact and international damage in recent decades and the greatest management strategy has been to avoid exposure to the virus. In Latin America, specifically in the Central American sub-region that includes seven independent countries: Guatemala, Belize, Honduras, El Salvador, Nicaragua, Costa Rica and Panama; the impact on the health system and specifically on primary care has been enormous. In Honduras and other Central American countries, the first cases reported were in larger and therefore most populous cities, which were affected in terms of reported cases. In early July 2020, a positivity of 41.1% was found, being the second highest in the region. (HENRIQUEZ-MARQUEZ *et al*, 2020).

Currently in Costa Rica, the lethality rate shows a wide variation of about 0.46%. At first sight, obvious correlations between accumulated incidences and parameters such as those related to climate, geographic characteristics or population peculiarities need to be studied when seroepidemiological surveys are made that allow a better approximation of the actual numbers of infected people. (CALLEJAS *et al*, 2020).

In mid-March, there was a substantial increase in cases of COVID-19, resulting in almost all countries in Latin America and the Caribbean (LAC). However, many areas throughout LAC, including Central America, have no information or have substantial gaps in information. Epidemiological details are often lacking due to the challenges in case-finding bias and the lack of serological tests. (ANDRUS *et al*, 2020). Although the implementation of strategies has been applied in most Latin American countries, there are intrinsic characteristics of the local community determined by many factors, such as demographic data, endemic infections and environmental conditions that can influence the outcome of infection in the region (BAUTISTA-MOLANO *et al*, 2020; YUAN, *et al*, 2020).

In Central America, Panama is the country most affected by the pandemic, followed by Honduras, Guatemala, El Salvador, Costa Rica and Nicaragua. At the global level, as the transmission of SARS-CoV-2 continues to advance, the main objective of health systems in many Latin American countries, and specifically in Central America, is the adoption of an epidemiological surveillance model adapted to the conditions of each country that reduce infection to a normal curve; The model is mainly based on the identification and screening of infected people, rapid response testing and treatment in critically injured patients, as well as the protection of people at greatest risk and vulnerability. In any case, the goal is to stop the explosive outbreak of the epidemic and accompany it with measures of isolation and confinement that, have been very drastic and severe in some countries and in others, looser and permissive (CALLEJAS *et al*, 2020).

Strict controls to limit entry and exit into American countries have also been an important impact strategy in containing the spread of COVID-19, as, with a large part of the region economically dependent on international tourism, measures regulating border flow balance competing demands for economic well-being and public health. It is expected that these control measures delay the spread of the virus and reduce the severity of the peak of the epidemic (MURPHY *et al*. 2020; ESCOBEDO *et al*, 2020).

While most Central American countries, as well as much of Asia and Africa are still preparing to detect and deal with COVID-19 outbreaks, it is essential to step up the training of the intercontinental and intracontinental health workforce, since in the Latin American region there is a great heterogeneity of political and social development, economic growth and political capacities (RODRIGUEZ-MORALES *et al*, 2020). Currently, even with the onset of vaccines, we cannot deny that in Latin America there are deficiencies in its health systems and infrastructure, especially a deficit of intensive care beds and mechanical ventilators necessary for the care of patients with severe infection, so that the risk of an overwhelming increase in deaths is latent (SÁNCHEZ-DUQUE *et al*, 2020).

The Pan American Health Organization works closely with central American and other continent countries to integrate COVID-19 into existing surveillance, based on surveillance of severe acute respiratory infections (hospitalized patients) and influenza-like diseases (outpatients), already in place as part of long-standing global influenza surveillance and more recently, for severe respiratory diseases since SARS and Influenza H1N1. This integration of approaches helps ensure long-term sustainable COVID-19 surveillance (ANDRUS *et al*., 2020).

As the SARS-CoV-2 pandemic progresses in Latin America, countries seek to acquire knowledge with other countries and implementing and following these methods is a challenge. As rich countries try new methods to maintain their economies, small island developing states face greater challenges. Islands belonging to the Dutch kingdom such as Curacao, St Martin and Aruba were quick to implement strict measures such as banning airspace and mitigation measures such as closing shops and restaurants. These measures, even though they were effective, caused great grief in the population because they generated the economic disaster because their economy depended on tourism and other sectors such as industry, which suffered from the lack of insum, causing 45% of the population to lose their jobs (some of these islands were already in economic recession), evidencing a worsening in an already weak economy (MARIA *et al*, 2020).

In the Caribbean region, for example, natural disasters, little investment in health and frequent arbovirus epidemics have been a constant problem. The COVID-19 outbreak occurred in March in the Dominican Republic, then Cuba and Puerto Rico and also on islands such as St Martin and Barbados and in countries such as Haiti, where only a handful of people understand that coronavirus contagion occurs from person to person, regardless of their religion or ethnicity, and most believe that the virus is God’s punishment, or that the elite manufactured the disease to annihilate minorities and the poorest, is the biggest of the problems encountered in virus mitigation (ESCOBEDO *et al*, 2020).

Latin America is marked by intense social inequality and with the arrival of SARS-CoV-2 this aspect was only more visible, demonstrating all the deficiency of the health system of these countries and its lack of infrastructure to deal with a pandemic. To correct these deficits, it is essential to rely on epidemiological surveillance, tracking new cases and establishing mandatory quarantine for those who come from outside, since according to all international organizations quarantine is the biggest step to reduce the rate (SÁNCHES-DUQUE, 2020).

Despite this, we at the Laboratory of Population Genetics and Computational Evolutionary Biology (LaBECom-UNIVISA) believe that contributing to genetic-population studies of SARS-CoV-2 in Central America can also be a valid strategy for virus mitigation, more precisely when we try to establish relationships and considerations about its molecular diversity. These considerations could help us in the support that, among other things, the more conserved the viral sequences found among all central American countries, the better the chances of resistance of the populations of these countries, since the genetic diversity of the entire Latin population may be reflected in the plasticity of clinical forms of COVID-19 found in these countries.

## 2. Objective

Test the levels of molecular diversity existing in complete Genomes of SARS-CoV-2 from Central America.

## 3. Methodology

### Database

The 38 complete SARS-CoV-2 genomes, publicly available on the National Biotechnology Information Center (NCBI) platform of the five Central American countries (Belize, Guatemala, Cuba, Jamaica, and Puerto Rico), were rescued from GENBANK (https://www.ncbi.nlm.nih.gov/labs/virus/vssi/#/virus?SeqType_s=Nucleotide&VirusLineage_ss=Severe%20acute%20respiratory%20syndrome%20coronavirus%202,%20taxid:2697049&Region_s=North%20America) on October 20, 2020.

### For tree construction using FIGTREE V 1.**4.4. (VLAD *et al*, 2008)**

To assemble the phylogenetic tree, we used the “Kimura 2-parameter” model with 100 pseudo-replications and as rates among sites we use the Gamma Distributed with Invariant Sites (G+I). The Newick tree, served as input for Figtree software.

### Genetic Structuring Analyses

The Analysis of Molecular Variance (AMOVA), Genetic Distance, mismatch, demographic and spatial expansion analyses, molecular diversity and evolutionary divergence time were obtained with the Software Arlequin v. 3.5 (EXCOFFIER *et al*., 2005) using 1000 random permutations (NEI and KUMAR, 2000).

**All steps of this process are described in**: https://dx.doi.org/10.17504/protocols.io.bmbvk2n6 (Félix *et al*., 2020).

## Results

Of the 38 sequences analyzed, with sizes ranging from 29,570 to 29,882 bp, only 79 sites remained preserved, revealing a high degree of polymorphism for the whole set. The graphical representation of these sites can be seen in a logo built with weblogo 3 software. (CROOKS *et al*., 2004), where the size of each nucleotide is proportional to its frequency for specific sites. (Figure 1 and Table 1).

**Table 1.** The *SARS-COV-2* genome sequences used in this study

| Accession | Release Date | Specie | Length | Geo Location | Host | Isolation Source | Collection Date |
| --- | --- | --- | --- | --- | --- | --- | --- |
| MT844023 | 05/08/2020 | SARS-CoV-2 | 29882 | Belize | *Homo sapiens* | - | 20/03/2020 |
| MT844024 | 05/08/2020 | SARS-CoV-2 | 29882 | Belize | *Homo sapiens* | - | 01/03/2020 |
| MT844026 | 05/08/2020 | SARS-CoV-2 | 29873 | Belize | *Homo sapiens* | - | 27/03/2020 |
| MT844027 | 05/08/2020 | SARS-CoV-2 | 29882 | Belize | *Homo sapiens* | - | 09/04/2020 |
| MT873896 | 11/08/2020 | SARS-CoV-2 | 29811 | Cuba | *Homo sapiens* | - | 19/03/2020 |
| MT873897 | 11/08/2020 | SARS-CoV-2 | 29800 | Cuba | *Homo sapiens* | - | 24/03/2020 |
| MT844029 | 05/08/2020 | SARS-CoV-2 | 29882 | Guatemala | *Homo sapiens* | oronasopharynx | 15/03/2020 |
| MT844031 | 05/08/2020 | SARS-CoV-2 | 29882 | Guatemala | *Homo sapiens* | oronasopharynx | 15/03/2020 |
| MT844032 | 05/08/2020 | SARS-CoV-2 | 29882 | Guatemala | *Homo sapiens* | oronasopharynx | 17/03/2020 |
| MT844033 | 05/08/2020 | SARS-CoV-2 | 29882 | Guatemala | *Homo sapiens* | oronasopharynx | 18/03/2020 |
| MT844034 | 05/08/2020 | SARS-CoV-2 | 29882 | Guatemala | *Homo sapiens* | oronasopharynx | 19/03/2020 |
| MT844036 | 05/08/2020 | SARS-CoV-2 | 29882 | Guatemala | *Homo sapiens* | oronasopharynx | 20/03/2020 |
| MT844038 | 05/08/2020 | SARS-CoV-2 | 29882 | Guatemala | *Homo sapiens* | oronasopharynx | 21/03/2020 |
| MT844047 | 05/08/2020 | *SARS-CoV-2* | 29882 | Guatemala | *Homo sapiens* | oronasopharynx | 13/03/2020 |
| MT844048 | 05/08/2020 | *SARS-CoV-2* | 29882 | Guatemala | *Homo sapiens* | oronasopharynx | 15/03/2020 |
| MT844049 | 05/08/2020 | *SARS-CoV-2* | 29882 | Guatemala | *Homo sapiens* | oronasopharynx | 15/03/2020 |
| MT507271 | 22/05/2020 | *SARS-CoV-2* | 29882 | Jamaica | *Homo sapiens* | swab | 09/03/2020 |
| MT507272 | 22/05/2020 | *SARS-CoV-2* | 29882 | Jamaica | *Homo sapiens* | swab | 11/03/2020 |
| MT507273 | 22/05/2020 | *SARS-CoV-2* | 29882 | Jamaica | *Homo sapiens* | swab | 14/03/2020 |
| MT507274 | 22/05/2020 | *SARS-CoV-2* | 29882 | Jamaica | *Homo sapiens* | swab | 16/03/2020 |
| MT507275 | 22/05/2020 | *SARS-CoV-2* | 29882 | Jamaica | *Homo sapiens* | swab | 16/03/2020 |
| MT507276 | 22/05/2020 | *SARS-CoV-2* | 29882 | Jamaica | *Homo sapiens* | swab | 17/03/2020 |
| MT507793 | 22/05/2020 | *SARS-CoV-2* | 29885 | Jamaica | *Homo sapiens* | swab | 11/03/2020 |
| MT507794 | 22/05/2020 | *SARS-CoV-2* | 29882 | Jamaica | *Homo sapiens* | swab | 16/03/2020 |
| MT419810 | 01/05/2020 | *SARS-CoV-2* | 29758 | Puerto Rico | *Homo sapiens* | oronasopharynx | 23/03/2020 |
| MT419811 | 01/05/2020 | *SARS-CoV-2* | 29743 | Puerto Rico | *Homo sapiens* | oronasopharynx | 23/03/2020 |
| MT419812 | 01/05/2020 | *SARS-CoV-2* | 29758 | Puerto Rico | *Homo sapiens* | oronasopharynx | 23/03/2020 |
| MT419813 | 01/05/2020 | *SARS-CoV-2* | 29570 | Puerto Rico | *Homo sapiens* | oronasopharynx | 23/03/2020 |
| MT419814 | 01/05/2020 | *SARS-CoV-2* | 29743 | Puerto Rico | *Homo sapiens* | oronasopharynx | 23/03/2020 |
| MT419815 | 01/05/2020 | *SARS-CoV-2* | 29743 | Puerto Rico | *Homo sapiens* | oronasopharynx | 24/03/2020 |
| MT419816 | 01/05/2020 | *SARS-CoV-2* | 29760 | Puerto Rico | *Homo sapiens* | oronasopharynx | 24/03/2020 |
| MT419817 | 01/05/2020 | *SARS-CoV-2* | 29570 | Puerto Rico | *Homo sapiens* | oronasopharynx | 23/03/2020 |
| MT419818 | 01/05/2020 | *SARS-CoV-2* | 29743 | Puerto Rico | *Homo sapiens* | oronasopharynx | 23/03/2020 |
| MT419819 | 01/05/2020 | *SARS-CoV-2* | 29743 | Puerto Rico | *Homo sapiens* | oronasopharynx | 24/03/2020 |
| MT419820 | 01/05/2020 | *SARS-CoV-2* | 29706 | Puerto Rico | *Homo sapiens* | oronasopharynx | 24/03/2020 |
| MT419821 | 01/05/2020 | *SARS-CoV-2* | 29743 | Puerto Rico | *Homo sapiens* | oronasopharynx | 01/04/2020 |
| MT419822 | 01/05/2020 | *SARS-CoV-2* | 29743 | Puerto Rico | *Homo sapiens* | oronasopharynx | 01/04/2020 |
| MW018448 | 18/09/2020 | *SARS-CoV-2* | 29825 | Puerto Rico | *Homo sapiens* | oronasopharynx | 06/07/2020 |

**Source:** NCBI Virus community portal

**Figure 1:**
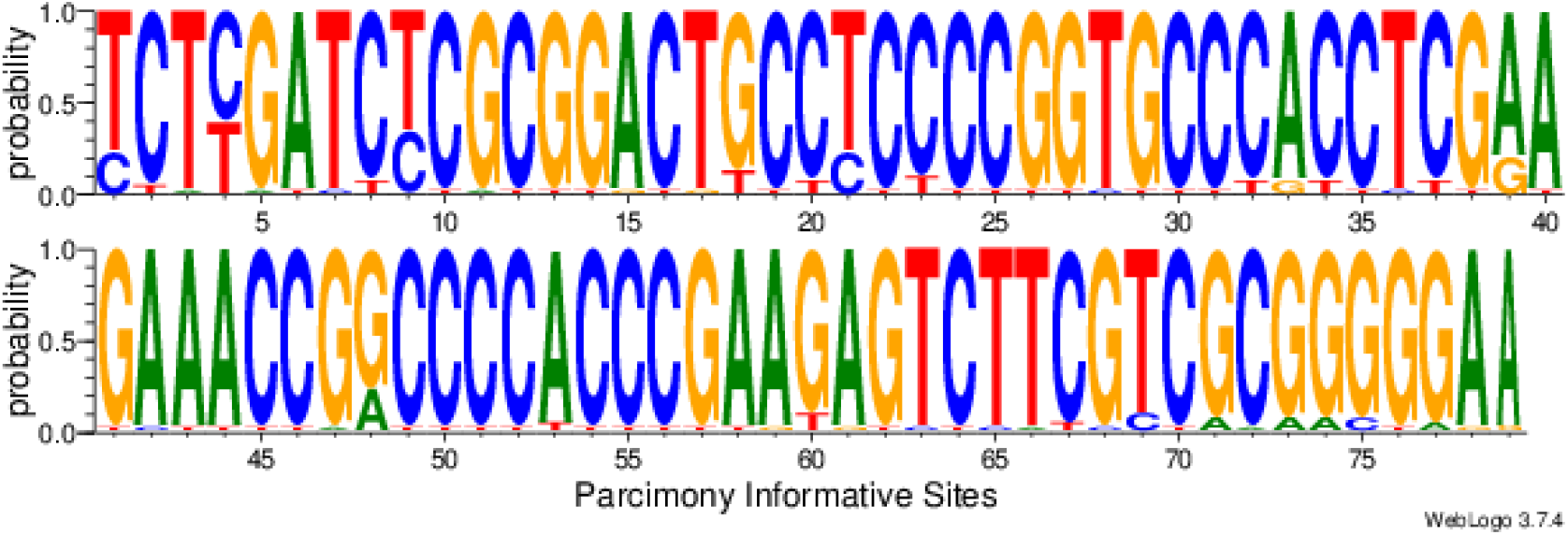
Graphic representation of 79 parsimonium-informative sites of complete Genomes of SARS-CoV-2 from Central America.

### Genetic Distance Analysis

Genetic distance and molecular variance (AMOVA) analyses were significant for the groups studied. The F_ST_ value (17%) pointed out the existence of a considerable diversity among the countries studied, with significant evolutionary divergences within and between the groups studied. These analyses indicate the countries of Guatemala and Jamaica as the most genetically distant and the countries of Cuba and Puerto Rico as the most similar. (Figure 2 and Table 2).

**Table 2.** Components of haplotypic variation and paired F_ST_ value for the 38 complete SARS-CoV-2 genomes from Central America.

| Source of variation | d.f. | Sum of squares | Variance components | Percentage of variation |
| --- | --- | --- | --- | --- |
| Among populations | 4 | 32.624 | 0.70557 $V_a$ | 17.15 |
| Within populations | 32 | 109.051 | 3.40785 $V_b$ | 82.85 |
| Total | 36 | 141.676 | 4.11342 |  |
| Fixation Index | $F_{ST}$: | **0.17153** | | |
| Significance tests (1023 permutations) |  |  |  |  |
| Va and $F_{ST}$: P (rand. value > obs. value) = 0.00000 | | | | |
| P (rand. value = obs. value) = 0.00000 |  |  |  |  |
| P-value = 0.00000+-0.00000 |  |  |  |  |

**Figure 2.**
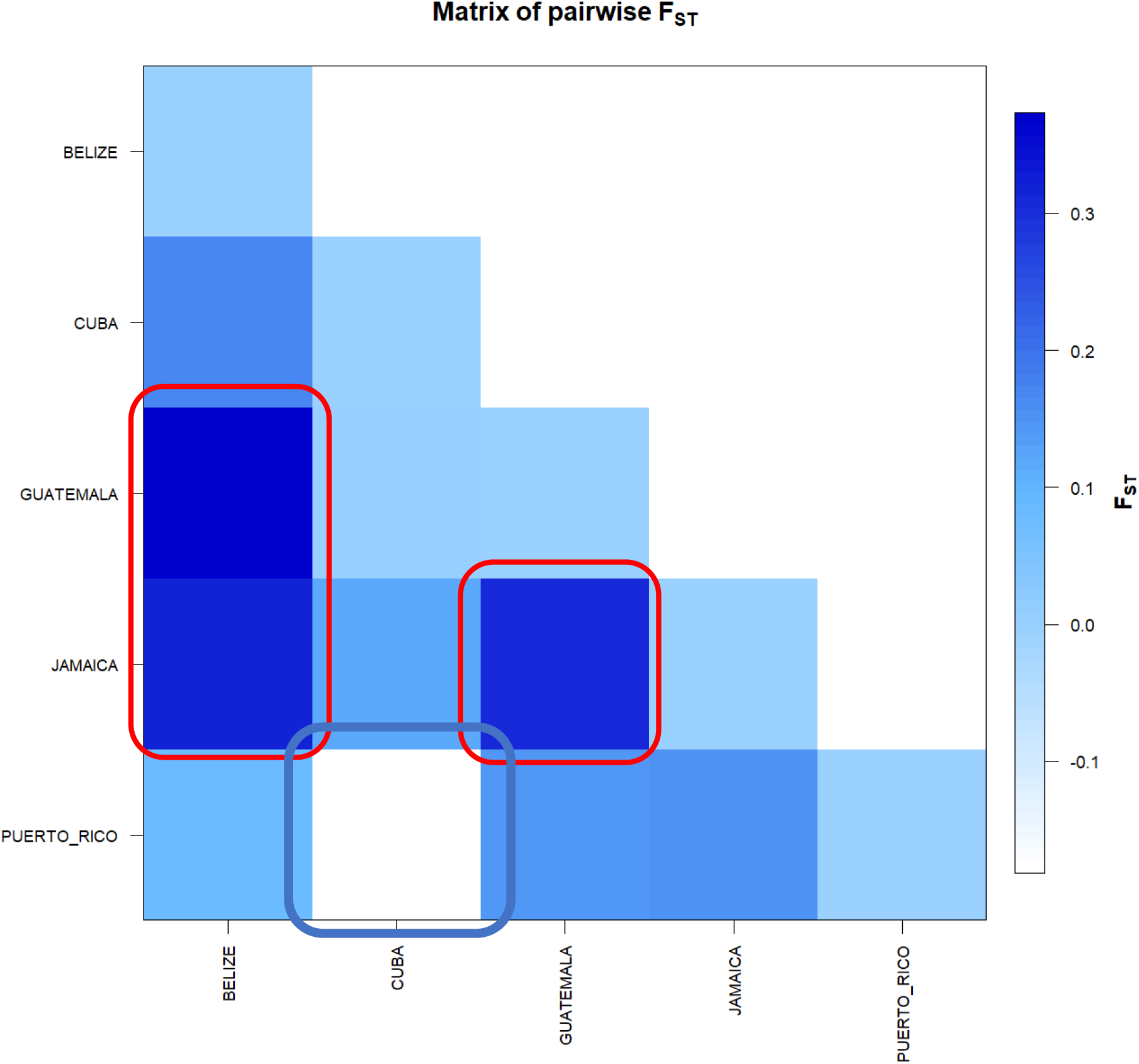
FST-based genetic distance matrix between for the 38 complete SARS-CoV-2 genomes from Central America. * Generated by the statistical package in R language using the output data of the Software Arlequin version 3.5.1.2.

The analyses confirmed the presence of high and statistically significant variations, evidencing a high genetic dissimilarity among all haplotypes. However, the use of the divergence matrix, in the construction of the tree helped in the recognition of minimal similarities between some haplotypes, including at different geographical points. The maximum divergence patterns were also obtained when less robust methods of phylogenetic pairing (e.g. UPGMA) were used, reflecting the non-haplotypic structure in the clades. With the use of the divergence matrix, it was possible to identify geographical variants that had less genetic distance and the “*a posteriori*” probabilities were able to separate the main clusters into additional small groups, confirming the presence of a minimum probability of kinship between haplotypes (Figure 3).

**Figure 3.**
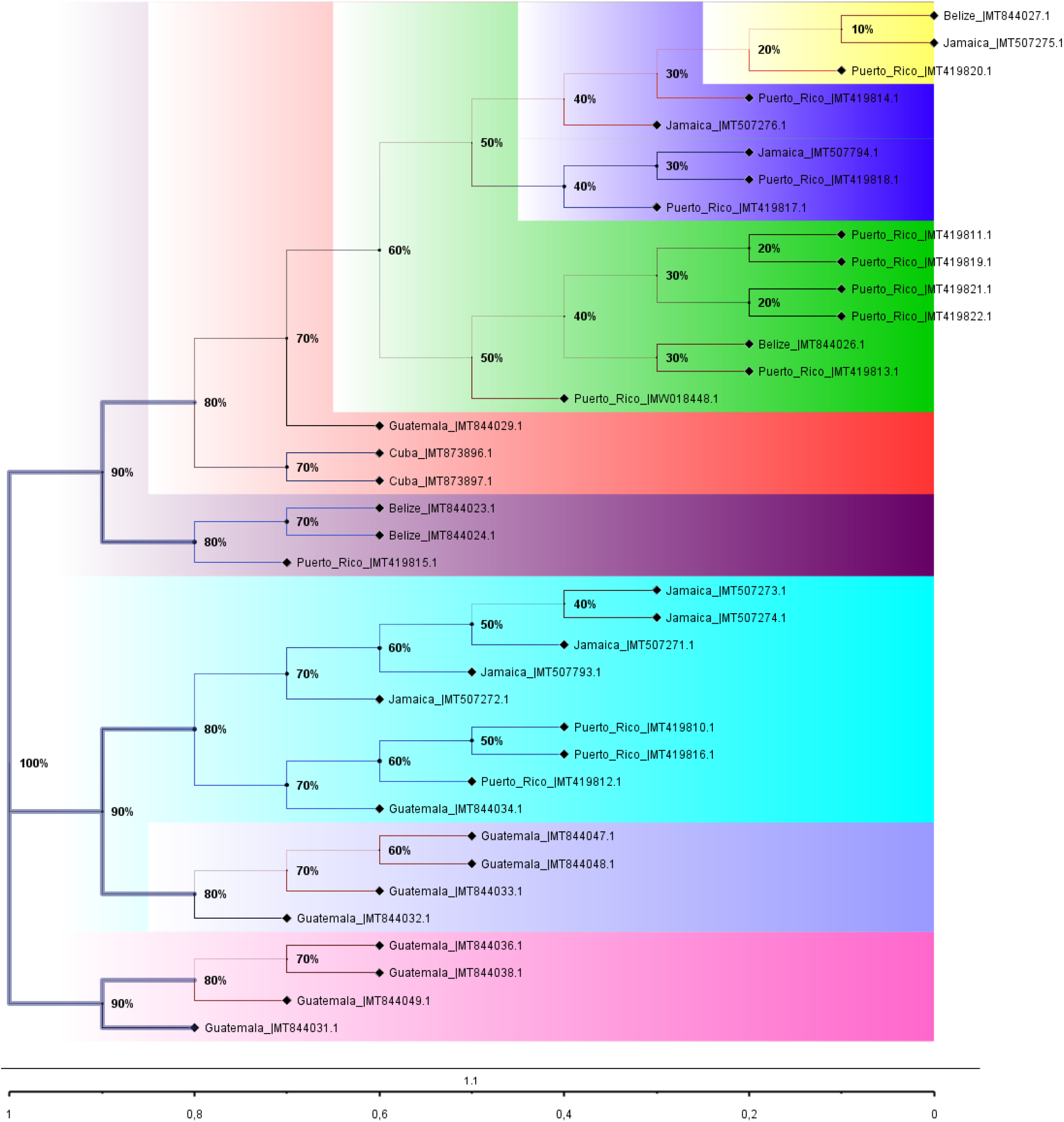
Phylogenetic Tree of the 38 complete Genomes of SARS-CoV-2 from Central America. We assume the need to display the “*a posteriori”* probabilities for each of the clades present, as well as the estimation of the age of each node. A reverse temporal scale was constructed to understand the evolutionary history of the set. We chose to draw the tree with thick lines and the clades were colored by selecting the branches. Finally, we export the tree in NEXUS and generate graphics files in PNG. * Tree built using FIGTREE V 1.4.4. (VLAD *et al*, 2008).

The *Tau* variations, related to the ancestry of some groups, revealed a significant time of divergence, supported by mismatch analysis and demographic and spatial expansion analyses (Table 3).

**Table 3.** Demographic and spatial expansion simulations based on the τ, θ, and M indices of sequences of the 38 complete Genomes of SARS-CoV-2 from Central America.

| Statistics | BELIZE | CUBA | GUATEMALA | JAMAICA | PUERTO RICO | Mean | s.d. |
| --- | --- | --- | --- | --- | --- | --- | --- |
| **Demographic expansion** |  |  |  |  |  |  |  |
| Tau | 8.00000 | 0.00000 | 1.50000 | 12.37307 | 3.00000 | 4.97461 | 5.11393 |
| Theta0 | 0.66250 | 0.00000 | 0.00000 | 0.00000 | 7.19998 | 1.57250 | 3.15891 |
| Theta1 | 3417.48520 | 0.00000 | 6829.37028 | 23.76217 | 6934.95629 | 3441.11479 | 3435.37960 |
| SSD | 0.42852 | 0.00000 | 0.04638 | 0.11977 | 0.01868 | 0.12267 | 0.17694 |
| Raggedness index | 1.00000 | 0.00000 | 0.15457 | 0.37628 | 0.04033 | 0.31424 | 0.41030 |
| **Spatial expansion** |  |  |  |  |  |  |  |
| Tau | 8.74785 | 0.00000 | 1.65237 | 12.03616 | 3.38650 | 5.16457 | 5.05541 |
| Theta | 0.00072 | 0.00000 | 0.01000 | 0.00072 | 7.78697 | 1.55968 | 3.48116 |
| M | 2.58419 | 0.00000 | 4397.92467 | 8.91409 | 4379.13801 | 1757.71219 | 2401.60969 |
| SSD | 0.34339 | 0.00000 | 0.04774 | 0.12062 | 0.01843 | 0.10603 | 0.14042 |
| Raggedness index | 1.00000 | 0.00000 | 0.15457 | 0.37628 | 0.04033 | 0.31424 | 0.41030 |

Molecular diversity analyses estimated by φ reflected a significant level of mutations among all haplotypes (transitions and transversions). The “indels” mutations (insertions or additions) were not found in any of the five groups studied. The Tajima and Fs de Fu tests showed disagreements between the estimates of general φ and π, but with negative and highly significant values, indicating, once again, the absence of population expansions. The irregularity index (R= Raggedness) with parametric bootstrap, simulated new values φ for before and after a supposed demographic expansion and, in this case, assumed a value equal to zero for all groups (Table 4); (Figure 4).

**Table 4.**
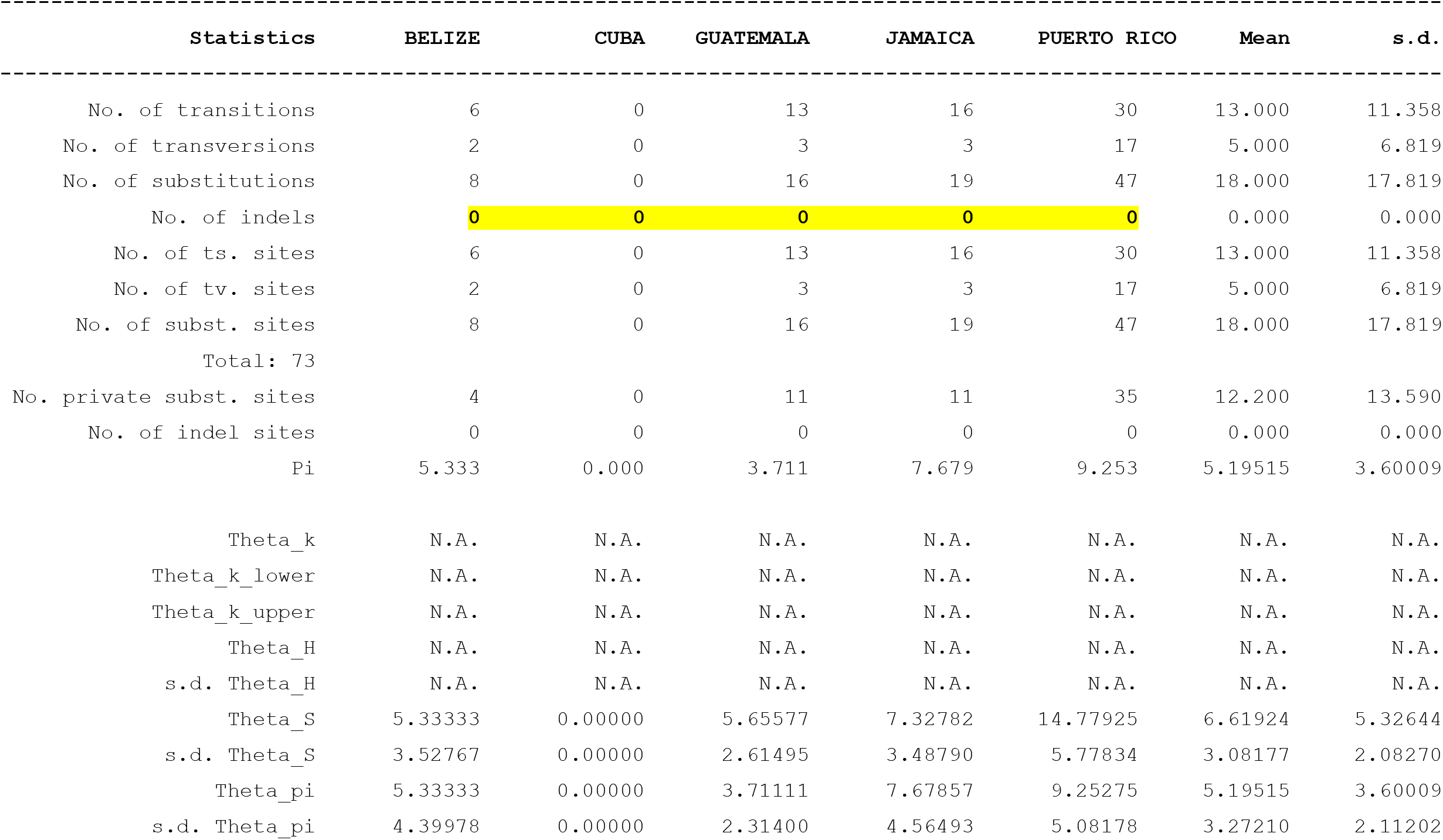
Molecular Diversity Indices of the 38 complete Genomes of SARS-CoV-2 from Central America.

**Figure 4.**
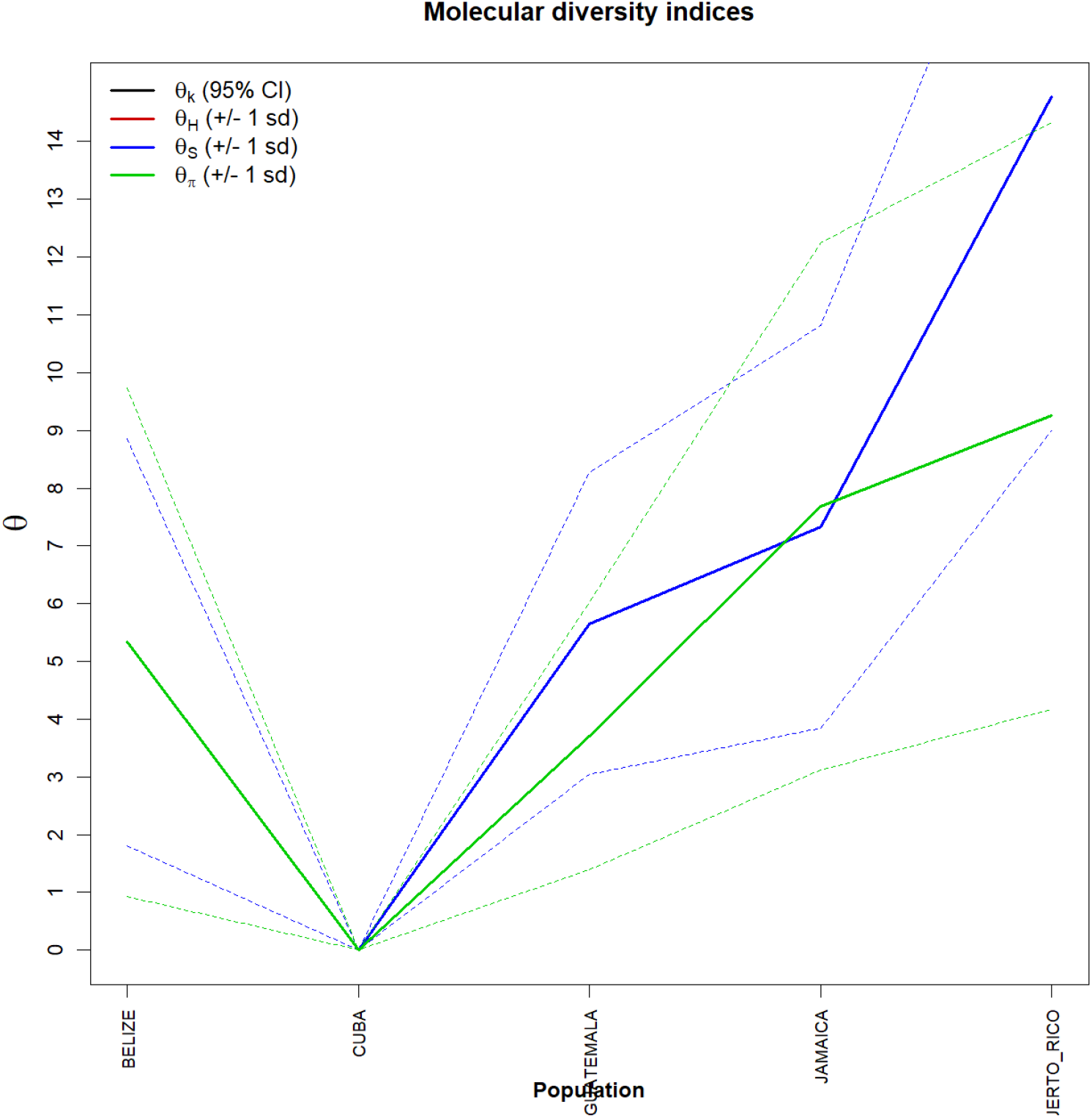
Graph of molecular diversity indices of the 38 complete Genomes of SARS-CoV-2 from Central America. In the graph the values of θ: (θk) Relationship between the expected number of alllos (k) and the sample size; (θH) Expected homozygosity in a balanced relationship between drift and mutation; (θS) Relationship between the number of segregating sites (S), sample size (n) and non-recombinant sites; (θπ) Relationship between the average number of paired differences (π) and θ. * Generated by the statistical package in R language using the output data of the Arlequin software version 3.5.1.2.

## 5. Discussion

With the use of analysis methodologies employed, it was possible to detect the existence of a small degree of similarity between the SARS-CoV-2 haplotypes of Central America. As no significant levels of structuring were found, we assumed that there were high levels of variation probably related to a gain of intermediate haplotypes over time, associated, perhaps, with a significant increase in genetic flow. The occurrence of geographical Isolation from past fragmentation events may have generated a continuous pattern of genetic divergence between the groups, since the high values found for genetic distance support the presence of a pattern of continuous divergence between haplotypes, as well as in the frequency of polymorphisms. This suggests that molecular diversity may be due to synonymous substitutions as the main components of variations. All analyses confirm that there is no consensus on the conservation of the SARS-CoV-2 genome in Central America, contrary to what was described by FELIX *et al*, 2020, for some countries in South America, and it is safe to state that the genetic variability of the Virus is different for subsets of countries throughout the American continent.

These considerations were also supported by simple phylogenetic pairing methodologies, such as UPGMA, which in this case, with a discontinuous pattern of genetic divergence between groups, showed a large number of branches with many mutational stages. These mutations possibly settled to drift due to the founder’s effect, which accompanies the dispersive behavior and/or loss of intermediate haplotypes throughout the generations. The values found for the genetic distance considered the minimum differences between the groups, as well as the inference of values greater than or equal to those observed in the proportion of these permutations, including the p-value of the test.

The discrimination of the five genetic entities was also perceived when the interhaplotypic variations were hierarchised in all components of covariance: by their intra- and inter-individual differences or by their intra- and intergroup differences, generating dendrograms that supported the idea that the significant differences found in the group of Guatemala, Jamaica and Puerto Rico, for example, can even be shared in their number, but not in their form, since the result of estimates of the average evolutionary divergence within the groups was low.

Since no relationship was made between genetic distance and geographic distance (mantel test) in this study, we assumed that the absence of genetic flow (observed by non-haplotypic sharing) between the studied regions is due to the presence of natural geographic barriers. The φ estimators, although extremely sensitive to any form of molecular variation, supported the uniformity between the results found by all the methodologies employed, and can be interpreted as a confirmation that there is no consensus in the conservation of sequences among the Central American countries studied, and it is safe to state that the large number of polymorphisms found should be directly reflected in a high variation for protein products of the virus, a fact that draws attention to the maintenance and increase of all public health actions employed.

## Data Availability

I declare that the SARS-CoV-2 genomes used in this study are publicly available on the National Biotechnology Information Center (NCBI) platform and that the protocols used in the methodology are our own and are also available at the link below.

https://www.ncbi.nlm.nih.gov/labs/virus/vssi/#/virus?SeqType_s=Nucleotide&VirusLineage_ss=Severe%20acute%20respiratory%20syndrome%20coronavirus%202,%20taxid:2697049&Region_s=North%20America

https://dx.doi.org/10.17504/protocols.io.bmbvk2n6

